# A systematic review of near-infrared spectroscopy in dementia

**DOI:** 10.1101/2022.11.23.22282361

**Authors:** Emilia Butters, Sruthi Srinivasan, John T O’Brien, Li Su, Gemma Bale

## Abstract

**Objectives:** This review aimed to evaluate previous studies using Near-infrared spectroscopy (NIRS) in dementia by summarising the results, determining the consensus in the literature, and delineating if, and how, NIRS experimental and analysis methods may be improved for future studies in dementia.

**Methods:** Three databases (PsychINFO, Medline, Embase) were searched for original research studies using NIRS in dementia and prodromal disease stages. We included both observational and randomised control trials, and studies published in English. Animal studies, conference abstracts, and reviews were excluded.

**Results:** From 759 identified records, 80 studies using NIRS in dementia and prodromal populations across a range of activation tasks testing memory (28), word retrieval (22), and motor (7) and visuo-spatial function (4), as well as in the resting state (29) were evaluated. Across these cognitive domains, dementia patients generally showed a blunted haemodynamic response, often localised to frontal regions of interest, and a lack of task-appropriate frontal lateralisation. Prodromal stages, such as Mild Cognitive Impairment, revealed mixed results and were associated with either diminished responses or hyperactivity, accompanied by reduced cognitive function, the latter suggesting a possible compensatory neural response which is not present at the dementia stage.

**Conclusion:** There is clear evidence of alterations in brain oxygenation in both dementia and prodromal stages across a range of cognitive domains and in the resting state, indicating an ability of NIRS to distinguish dementia from healthy ageing, or at-risk populations. A consensus as to the nature of these changes, however, is difficult to reach due to a lack of standardisation in optical techniques and processing methods. Further studies are required exploring more naturalistic settings and in a wider range of dementia subtypes.

## 1. Introduction

The most common form of dementia is Alzheimer’s Disease (AD), characterised by amyloid plaques, neurofibrillary tangles, memory impairment, cortical shrinking and hippocampal atrophy [1]. Other degenerative forms include Dementia with Lewy Bodies (DLB), characterised by Lewy body inclusions and motor symptoms, Fronto-Temporal Dementia (FTD), associated with fronto-temporal degeneration, and vascular dementia (VaD), caused by ischemia often due to stroke or atherosclerosis [1]. Dementia is a progressive disorder wherein the gradual cognitive decline in prodromal stages such as Mild Cognitive Impairment (MCI) is a critical target for early intervention. The identification of individuals at high-risk of developing dementia prior to irreversible damage is essential as although the conversion rate from MCI to AD is as high as 12% per year [2], around 16% of those with MCI revert to normal cognition within a year [3].

Advances in imaging and fluid biomarkers are rapidly progressing, however, there is a lack of brain-specific, low-cost, and readily accessible markers for clinical use. Imaging techniques such as Magnetic Resonance Imaging (MRI) and Positron Emission Tomography (PET) can be invasive, expensive, and limited to specialist centres. Conversely, fluid biomarkers do not provide regional brain information.

Near-infrared spectroscopy (NIRS) is a non-invasive neuroimaging technique which uses near-infrared light to measure brain oxygenation by exploiting the differing absorption spectra of absorbing molecules in the brain. Within an optical window of the near-infrared range (650-950 nm), oxygenated (HbO) and deoxygenated haemoglobin (HbR) are the primary absorbers of light. Consequently, in NIRS systems, two (or more) wavelengths of light are shone into the brain and the detected light attenuation is used to estimate the concentrations of HbO and HbR which, due to neurovascular coupling, are considered analogous to brain activity. In continuous-wave NIRS specifically, light attenuation due to absorption is indistinguishable from light attenuation due to scattering effects, making only *relative* concentration changes from baseline measurable.

NIRS is a low-cost, portable, and easy-to-use technology which can provide regional brain-specific measures of brain oxygenation and metabolism. Alongside the associations between dementia and vascular dysfunction, NIRS may have great potential for use in both dementia research and to support diagnosis and monitoring. The present article seeks to review the use of NIRS in dementia. The consensus in the literature shall be evaluated and future avenues for the adaptation of NIRS for use in dementia shall be delineated by critically evaluating the methodologies and analytical methods used.

## 2. Methods

### a. Search strategy

A systematic search of MEDLINE (1946 to 2021), Embase (1947 to 2021) and PsychINFO (1806 to 2021) was performed on the 26^th^ of September 2022 to identify relevant articles for inclusion. The search was conducted using the following search terms: (Cognitive impairment OR Cognitive disorder OR Cognitive decline OR Vascular dementia OR Cognitive dysfunction OR Neurocognitive disorder OR Alzheimer* OR Dement* OR AD OR FTD OR DLB OR LBD) AND (Near-infrared spectroscopy OR Near infrared spectroscopy OR NIRS OR oxyhaemoglobin OR Tissue oxygenation index). Additional studies were identified through cross-referencing the bibliographies of the included studies. Two authors (EB, SS) screened abstracts and titles for relevant articles using Covidence (Veritas Health Innovation Ltd., Australia). Conflicts were resolved by a third reviewer (GB). Full texts of the screened studies meeting the eligibility criteria were then evaluated for inclusion. PROSPERO CRD42021297315.

### b. Inclusion and exclusion criteria

Studies involving human subjects clinically diagnosed with dementia or in prodromal disease stages, and those published in English were included. Case-controlled studies testing both the target group and healthy controls, as well as those exclusively testing target groups, such as randomised control trials, were included. Studies not published in English, conference abstracts and animal studies were excluded.

### c. Data Extraction

Data was extracted by two reviewers (EB, SS) and stored in a data extraction form created in Microsoft Excel for Mac Version 16.42 (Microsoft Corporation, Redmond, Washington). The following information was extracted from the included studies: author, publication year, study design, sample size, and NIRS parameters and device.

### d. Quality assessment

The quality of the included studies was assessed using the Newcastle Ottawa scale [4] for case-controlled studies, the JADAD scale [5] for randomised control trials, and the National Heart, Lung, and Blood Institute quality assessment tool for observational cohort studies [6]. Quality assessment was performed by two authors (EB, SS). The results of this assessment are provided in the supplement (S2).

## 3. Results

The process for identifying relevant records is shown in Figure 1. The electronic search identified 759 records. Following title and abstract screening, 114 studies were eligible for full-text screening in which 6 studies were excluded due to the wrong patient population, 4 for wrong study design, 24 were conference proceedings or abstracts, and 1 was a book chapter. One study was also identified through cross-referencing making a total of 80 studies for final evaluation.

**Figure 1.**
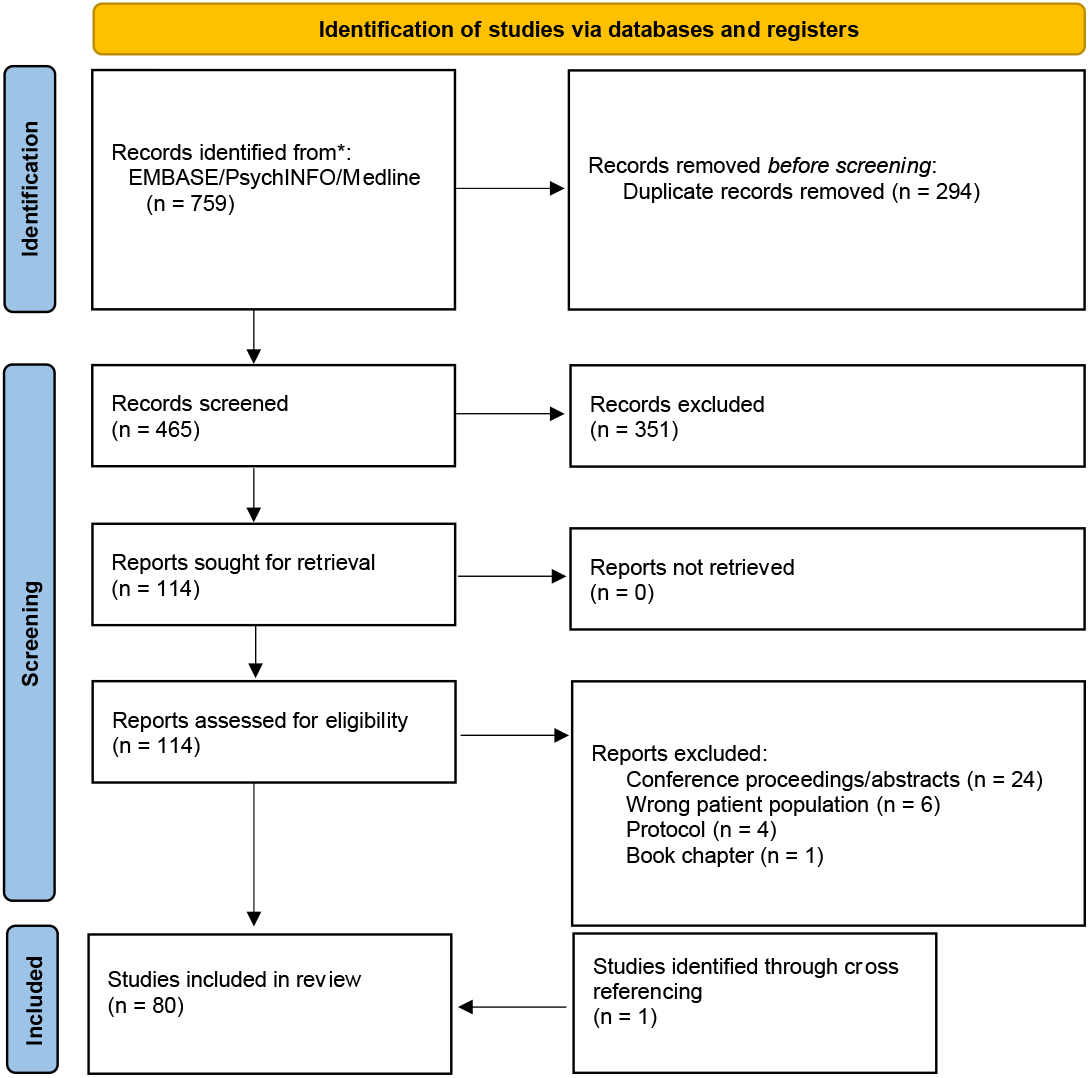
The PRISMA flow diagram [106] depicting the number of records at each stage of the selection process.

Since 1993, when the first paper was published using NIRS in dementia [7], there has been a steady increase in the number of papers published in the area (Figure 2). Of note is the gap in published studies between 1998 and 2004 which is likely due to a lack of commercially available NIRS systems for research.

**Figure 2.**
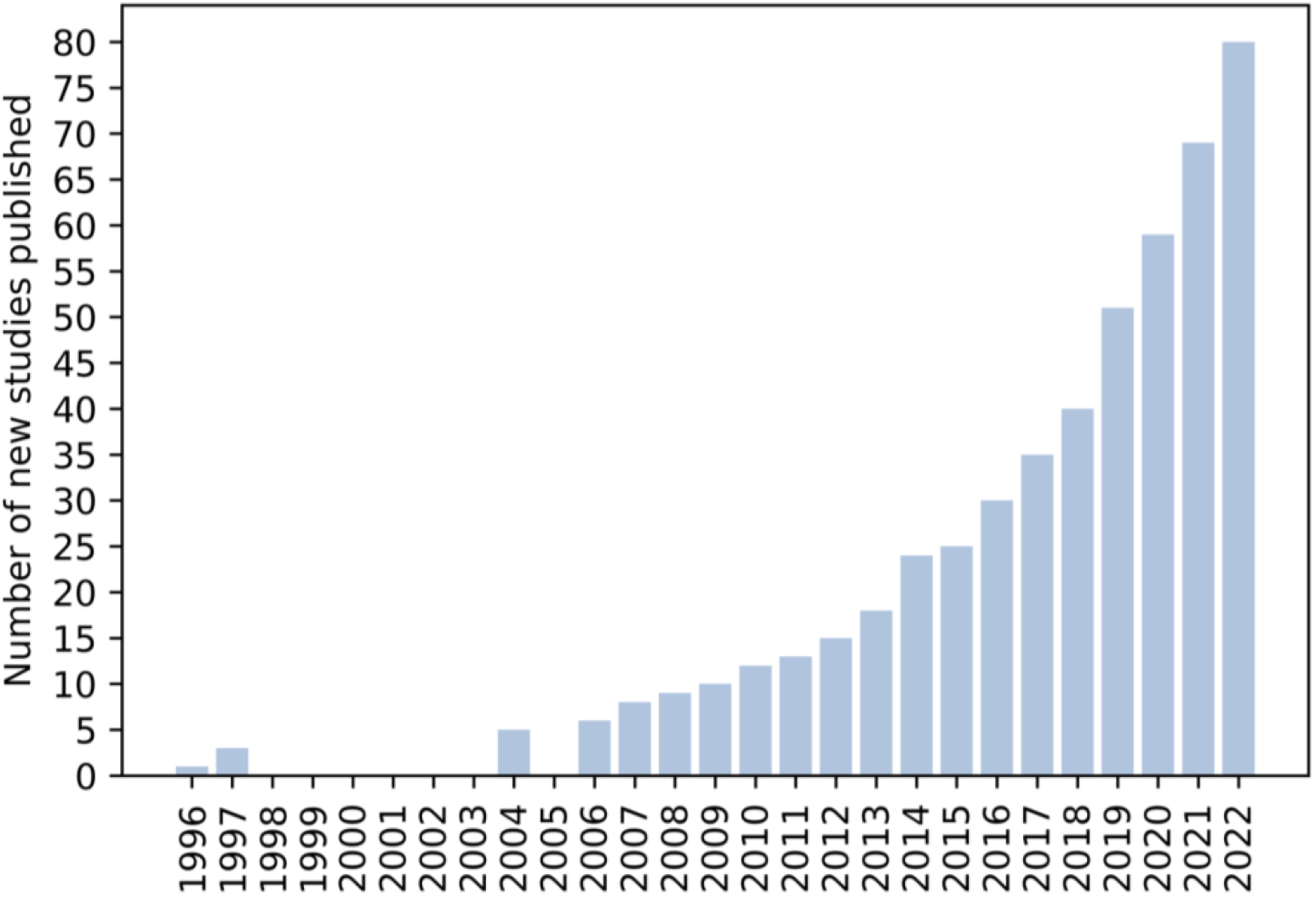
Cumulative histogram of the number of studies identified using NIRS in dementia.

The identified studies took several different approaches and covered six primary domains: resting state (29 studies, Table S1) and activation studies testing memory (28 studies, Table S2), word retrieval (22 studies, Table S3), motor (7 studies, Table S4) and visuo-spatial function (4 studies, Table S5), and other tasks such as oddball tasks (11 studies, Table S6).

## 4. Discussion

### a. Resting state measurements

#### Resting state brain oxygenation is reduced in prodromal dementia stages

A total of 29 studies explored resting-state brain oxygenation (Table S1, Figure 3a). Six employed a Tissue Oxygenation Index (TOI) [8]–[13]. This a commonly used metric in clinics which provides a measure of absolute tissue oxygen saturation, both arterial and venous, and is taken from a single measurement location. Several studies found reduced TOI in amnesic MCI (aMCI) [8], [11], and cognitively impaired individuals [12], compared to controls. In support of its clinical use, reduced TOI was associated with poorer MMSE [8] and memory scores [11] in aMCI. Conversely, TOI has been used as a marker of brain oxygenation to investigate therapeutic efficacy with mixed results. Two studies observed negligible TOI reactivity in AD with midazolam administration [14], [15], whereas Viola et al [13] observed significant increases in TOI in AD with brain reperfusion rehabilitation therapy alongside improved MMSE scores. The unclear nature of the alterations in TOI may be due to issues with intra-device variation [16]. With regards to the use of TOI outside of the clinic, as it only provides a single spatially invariant measure, it does not capture spatial variations in brain oxygenation.

**Figure 3.**
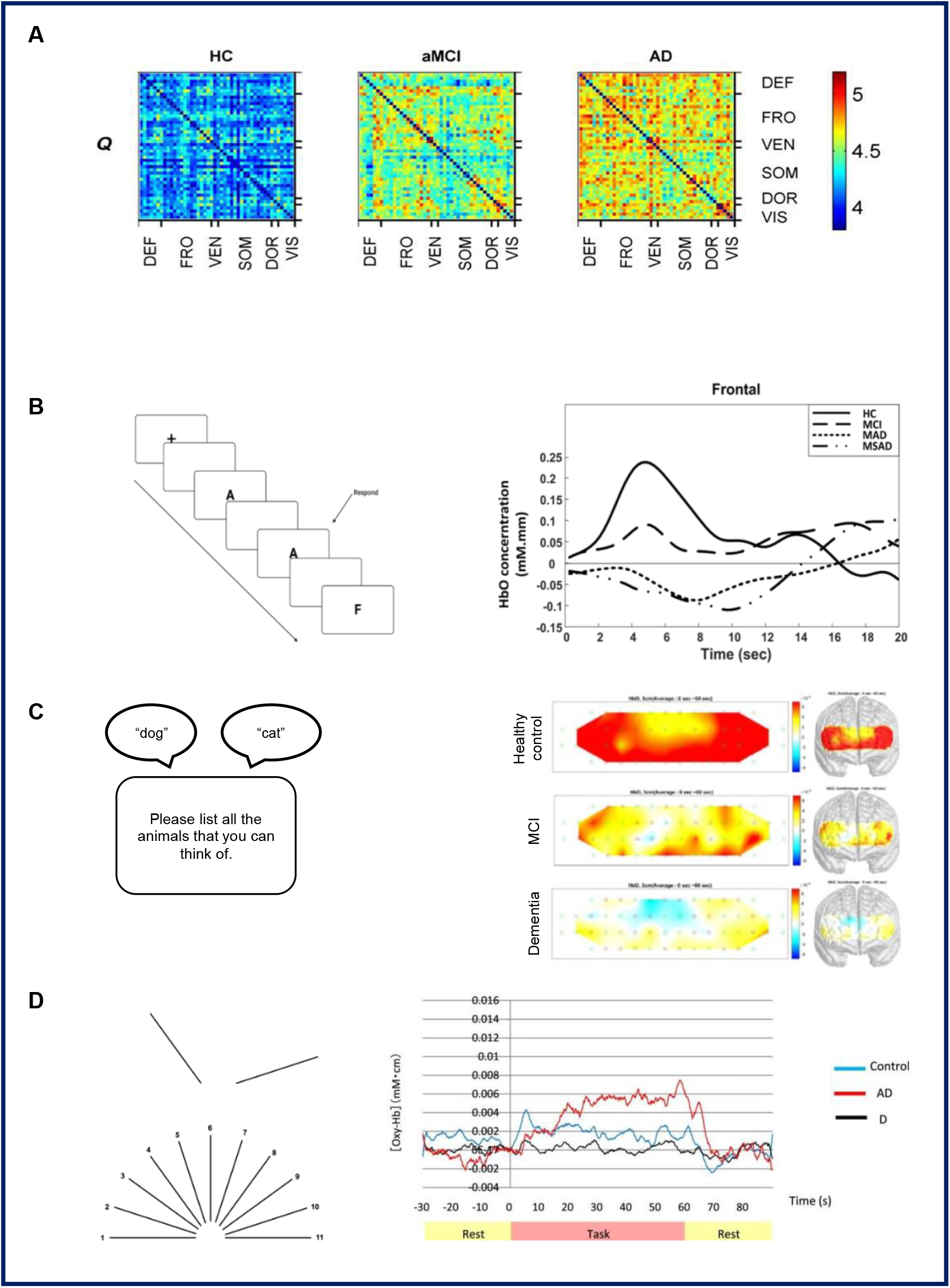
Increased resting-state functional connectivity variability (Q) from healthy controls to mild cognitive impairment, to Alzheimer’s Disease [34].. **(B)** *Left*. Schematic overview of the 1-back task. *Right*. Mild cognitive impairment is associated with a reduced and delayed rise in the haemodynamic response and Alzheimer’s Disease is associated with a decrease and delay in the response compared to healthy controls during memory encoding [49]. **(C)** *Left*. Schematic representation of the semantic verbal fluency task. *Right* Healthy controls have higher pre-frontal cortex activity compared to both mild cognitive impairment and dementia groups [27]. **(D)** *Left*. An example of the Benton Line Orientation task [89]. *Right*. Significantly reduced average HbO change in individuals with late life depression compared to Alzheimer’s Disease in a parietal channel during a visuo-spatial task [74].

Another commonly used method to measure resting state oxygenation, or rather *cerebrovascular reactivity* (i.e., the increase in HbO during rapid vasodilation caused by CO_2_ inhalation), is through sit-stand manoeuvres or CO_2_ challenges which yielded mixed results as to differences between dementia and MCI, and controls [17]–[19]. Interestingly resting state oxygenation during such challenges reflects oxygenation increases caused by acupuncture therapy and galantamine treatment in MCI [20], VaD [21], [22], and AD [23]. In contrast, purely resting state data does not appear to differentiate between AD [24]–[26] and MCI [27], and controls.

A pathophysiological process of interest in dementia is neurovascular decoupling [28] which can be explored using a multi-modal approach. Only two studies explored neurovascular decoupling in AD [26] and aMCI [19] using EEG-NIRS, however, these were limited by a lack of subject-specific information or low channel counts, preventing the exploration of spatial patterns of decoupling. Additionally, using broadband NIRS to measure neurometabolism would be invaluable to understand neurovascular coupling but has not been used in dementia.

#### Disorganisation of the cortex in dementia was identified via several computational methods

Many studies explored connectivity, several of which identified disturbances, the nature of which is not well defined. This is part due to the diverse methods and metrics used to quantify connectivity across studies. One such method is ‘effective connectivity’, i.e., the causal influence of one region over another, which is reduced in MCI across several regions such as the bilateral prefrontal cortex (PFC), in which stronger coupling was associated with improved cognitive scores [29]. Alternatively, the signal time courses can be correlated to calculate correlation coefficients. Reduced coefficients were observed in the PFC in AD [30] with greater HbR connectivity in the right hemisphere in MCI [31]. Taken together these results support the hypothesis for a compensatory response in prodromal stages to support declining cognitive function which fails in dementia stages [32], although the diagnostic relevance of the HbR signal is unclear [33].

With regards to specific regions of interest, both MCI and AD show disturbances in dynamic functional connectivity (i.e. accounting for the temporal variability of connectivity) within long-distance connections in prefrontal and parietal cortices, and in the Default Mode Network (DMN) and fronto-parietal networks [34] (Fig 3b). Another method is calculating the ‘entropy’, i.e., complexity, of a signal, which is considered to reflect cognitive ability. Entropy has been observed to be reduced in AD, which in accordance with Niu et al. [34], was localised to regions in the DMN, fronto-parietal and ventral/dorsal attention networks [35].

The application of another powerful computational method, machine learning (ML), to NIRS has been growing rapidly in popularity within the healthcare sector. Despite this, few studies (10) used ML analyses, and only one focused on the prediction of a continuous variable [36] while the rest focused on classification of dementia stage. Most used simple ML models such as support vector machines and linear discriminant analysis. This is notable as recent studies have demonstrated higher dementia classification accuracies using more complex ML or deep learning models [24] in comparison to traditional models. Two studies performed classification on resting state data, finding that classification of MCI from controls was more accurate using HbO compared to HbR [33]. The only study identified using broadband NIRS classified AD, MCI, and controls from their optical spectrum, finding a feature at 895 nm to be best at differentiating between AD and MCI [37], although what this indicates is unclear as the biological substance contributing to this peak could not be identified by the authors.

Only a few included studies (3) focused on multi-class classification, while the rest focus on binary classification between MCI/AD and controls. Chiarelli et al [26] used estimates of neurovascular coupling strength calculated from simultaneous EEG-fNIRS recordings and a multivariate linear regression approach to perform binary classification of AD and controls. In line with Cicalese et al [38] which performed multi-class classification, classification accuracies were improved when using combined EEG-fNIRS features [26].

Most discussed studies are limited by small group sizes and group imbalance, which do not provide enough training examples per group for a sufficiently robust model. With larger volumes of patient data, prediction, and finer-scale, classification tasks can be realised with high accuracies. Finally, none of the included studies focus on interpretable ML, such that none of the features used in making a final prediction can be interpreted in a clinical context.

### b. Functional measurements

#### Evidence for both hypo- and hyper-activation in dementia and prodromal groups during memory tasks

Overall, 28 studies explored memory function in dementia (Table S2), many of which used the n-back task to test memory function (13), with mixed results. This task evaluates working memory (WM) and frontal regions, making it good for use with NIRS as it avoids monitoring through hair. Subjects are presented with a sequence of letters and must indicate whether the presented letter was the same as that just before (one-back) or before last (two-back). Two studies observed blunted haemodynamic responses in MCI [39], [40], with a gradation from healthy controls to MCI to AD [24], whereas three found no difference in functional response [27], [41] or connectivity [31] between MCI and controls. Interestingly, one study identified hyperactivation in MCI compared to controls [42]. Perhaps the discriminatory ability of the n-back task is more subtle: there is evidence for differential WM load modulation across disease stages. Some observed differences in activation between MCI and controls only with high WM loads [43], [44] and others identified WM load modulation only in controls [45], [46].

With respect to the use of WM tasks in the clinic, most studies reporting correlation results identified positive correlations between the magnitude of the HbO signal or functional connectivity metrics [47], and behavioural or clinical scores [44], [48]–[51] (Fig 3b) such that greater oxygenation was associated with better scores. Encouragingly, haemodynamic responses to the n-back task have been validated as having strong diagnostic potential using convolutional neural networks [39], [43]. Responses to the n-back task may also be sensitive markers for therapy responses, demonstrated by increases in oxygenation [20], [48], [52]. Perhaps surprisingly, two studies found improved memory performance to be associated with *reductions* in frontal activation in MCI with photobiomodulation therapy [53] and VR-based physical/cognitive training [54]. This may further support the idea of a compensatory hyperactive response in prodromal stages.

Evidence for this also comes from studies using other WM tasks, such as the delayed matching to sample task or the digit verbal span task. Higher connectivity in aMCI [47], and larger HbO changes [55], [56] and greater entropy [57] have been observed in AD [57] and cognitive decline [55] compared to controls. However, similarly to the n-back task, several studies also identified hypoactivation in AD [20], [35], [58], [59], MCI [52], and specifically in aMCI [47], [50], [51]. Such variable responses are likely due to differences in analysis e.g., graph theory vs. entropy, task design, and the range of signal metrics used.

#### Word retrieval is associated with blunted haemodynamic responses and distinguishable patterns of activation across dementia type and stage

All the included studies assessing word retrieval (22, Table S3) used the verbal fluency task (VFT), or a modification of such. This is a frequently used paradigm in dementia in which subjects must words within a given category (‘semantic’) or beginning with a specific letter (‘phonemic’). Patient groups generally performed worse than controls, as was the case for AD [60]–[63], MCI [31], [61], [62], [64], asymptomatic AD [24], and the behavioural variant of FTD [62]. Such reduced behavioural performance was accompanied by smaller haemodynamic responses in AD [65]–[69], characterised by a longer latency [61], smaller peak amplitude, smaller area under the waveform, and lower amplitude [68]. These results largely agree with those from other imaging modalities, including hypometabolism identified using PET [70], an overall ‘slowing’ of neocortical EEG characterised by deactivated, synchronised patterns of activity [71], and altered functional connectivity using fMRI [72].

Several studies specifically focused on MCI, with varied results. A few studies observed hypoactivation [27], [40], [43], [73] (Fig 3c), particularly in the right parietal region [66], and reduced inter-hemispheric connectivity [31]. However, upon classifying between MCI and healthy controls using the HbO signal, the VFT was not as stable an indicator of MCI as the n-back task [39]. In support of a lack of diagnostic potential of the VFT, Baik et al. [27] did not identify differences between MCI and AD. In line with this, the association between the magnitude of the haemodynamic response and clinically-relevant features such as MMSE score [66], [68], [74], or behavioural performance [62], [65], [75], [76], is unclear.

Although the magnitude of the haemodynamic response during the VFT may not be clinically useful, the spatial patterns of activation during the VFT may differentiate between healthy ageing and dementia, as well as across MCI type [41], [64]. For example, differences between AD and controls are localised to frontal and bilateral parietal regions [67], whereas differences between AD and MCI are localised to right parietal regions [66]. A loss of activation asymmetry is also evident in both dementia [63], [65] and MCI [64] during word retrieval, however, one study found no significant lateralisation in either controls or MCI [73]. Despite this, the extent of lateralisation has been suggested to be a potential biomarker which indicates the recruitment of contralateral resources to support declining function [64], as is supported by the fMRI literature [77].

#### Motor tasks previously used in NIRS may be too simplistic

Of the seven studies testing motor function (Table S4), five used the dual-task walking paradigm [78]–[82] with wearable NIRS devices. This paradigm involves performing a single task (e.g., walking on their own) and a dual task, (e.g., completing a cognitive task, such as a VFT, whilst walking). This task revealed a potentially non-linear relationship between dementia severity, cerebral oxygenation, and motor performance, unlike memory function [24] or word retrieval [61]. For example, people with memory complaints had higher activation during dual-task walking compared to controls, whereas people with dementia had higher activation compared to both healthy controls and people with memory complaints in single-task walking, yet significantly reduced activation in dual-task walking [79]. In addition, certain studies found differences between single- and dual-walking in MCI [78] or associations between cognitive scores and activation [80], whereas others did not [81]. Such a lack of consensus is likely due to the fact that all of these studies, except one, recorded from the frontal cortex, which has a large degree of inter-subject variability [83], as opposed to recording from the motor cortex.

Concerning studies directly assessing motor function, none used naturalistic tasks, such as walking or social interaction, but used simplistic motor tasks, such as hand-grip movements [84] and finger tapping [85]. This is surprising due to the advantages that NIRS has in terms of low sensitivity to movement and lack of physical restrictions, and the characteristic motor symptoms of certain dementia subtypes, such as DLB [86], which cannot be easily explored using imaging techniques such as MRI or PET. However, the emergence of wearable NIRS is relatively recent which may explain the lack of naturalistic task designs.

#### More demanding visuo-spatial tasks may reveal clearer deficits in dementia

Four studies explored visuo-spatial processing [25], [74], [87], [88] (Table S5, Fig 3d). Three of these used angle discrimination tasks, such as the Benton Line Orientation task, which requires participants to judge the angle at which a presented line is oriented [89]. However, these yielded unclear or contradictory alterations in patient groups [25], [74], [88], possibly due to a lack of standardised methodologies making it difficult to compare across studies. For example, Zeller et al [25] use a combined ‘dementia’ patient group. The absence of performance difference across groups [25], [74] may also suggest that more demanding visuo-spatial tasks are required to reveal differences in the NIRS data.

#### A handful of studies used ‘unconventional’ stimuli or oddball tasks

Three studies explored sensory responses in dementia using NIRS (Table S6), such as responses to music [90] and olfactory stimuli, both of which discriminated healthy ageing from prodromal [91] and dementia stages [92].

Alternatively, seven studies employed oddball tasks such as the Stroop task. Three found no difference in the haemodynamic response [27], [43] and connectivity [31] between MCI and controls, whereas three found reduced frontal activation in MCI [39], [40] and AD [24]. As most of these studies used the same task design and patient groups, except for Ho et al [24] which used a four-minute task block, these mixed results may be surprising. This could be due to differences in statistical methods and signal metrics used to determine activation.

### c. Experimental methods

#### A wide range of NIRS devices and analysis methods were used which may underlie the range of results observed across studies

All of the included studies used continuous-wave NIRS systems bar [36] which used a time-resolved system, and [26] which used a frequency-domain system. Concerning analysis methods, none of the included studies performed subject-specific image reconstruction or source localisation. The variable results of the included studies may ultimately be explained by the widespread cortical atrophy and shrinkage present in dementia [93]. The subsequent increased distance of the cortex from the scalp may lead to data being recorded from extracerebral tissues rather than from the cortex. The integration of subject-specific anatomical data is thus necessary to avoid apparent differences in function being caused by anatomical variability or structural degeneration. High-density, variable length NIRS channel systems may also achieve better sensitivity to the cortex in dementia [94].

Such subject-specific information is also necessary for creating detailed topographical maps of brain activity using High-density Diffuse Optical Tomography (HD-DOT). Whilst no studies used HD-DOT and only a few studies used high-density systems (e.g., [27], [42]), Talamonti et al [82] used DOT and Li et al [59] performed source localisation of the NIRS signal, however, neither of these used subject-specific information. Moreover, many studies only analysed the HbO signal and discarded the HbR signal, citing a higher signal-to-noise ratio of the HbO signal and greater correlation with BOLD fMRI signal [95]. Although the individual diagnostic potential of the HbO and HbR signal is not clear [33], several studies do find significant differences between these two signals (e.g., [96]), with HbR possibly being more relevant than HbO [73].

Almost half of the reviewed studies either do not describe or have not performed motion correction. Older papers (1990s-2000s) usually did not correct for motion as such methods are a more recent development, however, many newer papers did not do so either. Instead, many of these studies state that participants were instructed to remain still during recording (e.g., [13]) or that cables and optodes were tightly fixed to prevent motion (e.g., [17], [23]). Some performed motion detection and excluded channels or blocks that exhibited sharp signal changes (e.g., [24], [27]). Others performed explicit motion correction using a variety of algorithms, the most common of which are cubic spline interpolation (e.g., [18]) and wavelet-based artifact removal (e.g., [57]). Similarly, several studies did not perform short-channel regression (e.g., [29]) to remove the influence of systematic scalp data. Most studies also only record from pre-specified regions of interest, limiting functional connectivity analyses. A more comprehensive review of the optical methods and their use in dementia is available at Srinivasan et al. (under review in Neurophotonics).

#### The majority of studies focused on AD, with few studies exploring less common types of dementia

A large proportion of studies (33) focused on AD, with 47 in MCI and only 3 in VaD, 1 in FTD, and none in DLB. The mixed evidence observed in MCI is perhaps due to the relatively small number of studies (11) that directly compare MCI with AD, and as MCI can be difficult to diagnose and classify into subtypes [97]. There was only a single longitudinal study exploring how brain oxygenation changes with disease progression [82], in which exploring even earlier prodromal stages such as APOE-4 carriers [98] is necessary for the assessment of NIRS’ clinical value. Few studies used NIRS simultaneously with other imaging modalities: 1 PET, 3 EEG, and 1 fMRI. Additionally, many, particularly those measuring task-related activation, recorded exclusively from frontal regions. This is despite the established posterior degeneration in AD and DLB [99], areas not generally recorded from in the included studies.

## 5. Conclusion

Broadly, the previous literature identified disorganisation of the cortex, involving the DMN, fronto-parietal networks, and long-range connections across the brain (e.g., [34]). Dementia presented with hypoactivation (e.g., [35], [50]) with strong evidence for a generally suppressed haemodynamic response across cognitive domains. In prodromal stages, several studies found hypoactivation [41], [66] whereas others identified a possible compensatory response in the form of hyperactivation [42], [61]. Alongside the generally blunted haemodynamic response in dementia, these mixed findings partially agree with the hypothesis of a ‘break point’ in prodromal stages [100]. This review highlights the necessity for standardised protocols both with regards to experimental design, e.g., ecologically valid task designs, and analysis methods, e.g., using subject-specific structural information for source localisation, for more holistic and generalisable outcomes.

## Data Availability

All data in the present work is available online.

## 6. Acknowledgements

EB, GB, and SS are supported by the Giana Angelopoulos Programme for Science Technology and Innovation. JOB and LS are supported by the Cambridge NIHR Biomedical Research Centre and the Cambridge Centre for Parkinson’s Plus Disorders.

