## Supplementary material for "A systematic review of near-infrared spectroscopy in dementia"

### 8. Supplement

a. S1: Characteristics of the included studies

**Table S1.** Characteristics of the included studies reporting resting-state near-infrared spectroscopy data in dementia and prodromal stages.

| **First author, year** | **Participant characteristics (*n*)** | **Task design** | **NIRS channels (*n* sources, detectors)** | **NIRS device** | **Main findings** |
|---|---|---|---|---|---|
| Babiloni, 2014 [19] | aMCI (10), controls (10) | Resting state during $CO_2$ challenge | 2 frontal channels (8S, 2D) | ISS Oximeter (Iss Inc.; IL, USA) | HbO and HbR time courses and amplitudes were similar between groups |
| Baik, 2021 [27] * | MCI (22), dementia (20), controls (18) | Resting state | 48 frontal channels (24S, 32D) | NIRSIT (OEBLAB Inc.; Seoul, Korea) | No difference in PFC activity between the groups.<br>No correlation between PFC activity and MMSE/MoCA scores. |
| Bar, 2007 [21] | AD (17), VaD (17), controls (20) | Resting state during $CO_2$ challenge, before/after galantamine treatment | 1 left frontal channel (1S, 1D) | NIRO 500 (Hamamatsu Photonics CO.; Hamamatsu, Japan) | Reduced absolute HbO in VaD compared to young and elderly controls, and in AD compared to young controls<br>Increased absolute HbO after galantamine treatment in VaD.<br>Correlation between MMSE and absolute HbO in VaD and AD. |
| Bu, 2019 [29] | MCI (26), controls (28) | Resting state | 14 channels across frontal, motor, and occipital cortex (10S, 8D) | Nirsmart (Danyang Huichuang Medical Equipment Co, Ltd.; Shaanxi, China) | Effective connectivity between ROIs lower in MCI compared to controls. |
| Canario, 2022 [101] | MCI (29), AD (25), controls (34) | Resting state | 46 whole-head channels (12S, 24D) | CW6 (TechEn Inc.; MA, USA) | No significant difference in functional connectivity between groups.<br>Increased segregation and hemispheric connectivity differences in AD as compared to MCI and controls. |
| Chiarelli, 2021 [26] | AD (17), controls (18) | Resting state | 16 frontal channels (32S, 4D) | Imagent (ISS Inc.; IL, USA) | No difference in mean HbO or HbR signal between AD and controls.<br>Significantly reduced coupling strength in AD compared to controls. |
| Fallgatter, 1997 [63] | AD (10), controls (10) | Resting state (prestimulus baseline) | 2 frontal channels (2S, 4D) | Critikon 2020 Cerebral Redox Monitor (Johnson and Johnson Medical) | Reduced absolute HbO in AD compared to controls.<br>Higher absolute HbO and Hb in the left compared to the right hemisphere for both AD and controls. |
| Ghafoor, 2019 [20] * | MCI (11), controls (11) | Resting state, before/after acupuncture therapy | 20 frontal channels (8S, 7D) | NIRScout (NIRx Medizintechnik GmbH; Berlin, Germany) | Lower mean global network and nodal efficiency in MCI compared to controls<br>Increased functional connectivity in MCI with acupuncture treatment. |
| Greco, 2021 [37] | AD (20), MCI (12), controls (13) | Resting state | 1 frontal channel (1S, 1D) | In house device. | Spectral feature at 895nm which distinguished AD from MCI, the biological identity of which was not identified. |

| | | | | | |
|---|---|---|---|---|---|
| Ho, 2022 [24] * | Prodromal AD (50), AD dementia (9), asymptomatic AD (28), controls (53) | Resting state | 6 frontal channels (2S, 5D) | In house device. | No significant difference in HbO concentration change between groups. |
| Li, 2018 [35] | aMCI (27), AD (24), controls (31) | Resting state | 46 whole-head channels (12S, 24D) | CW6 (TechEn Inc.; MA, USA) | Reduced brain signal complexity of HbO in AD compared to controls.<br>Decreased complexity in default mode, frontoparietal, dorsal and ventral attention networks in AD compared to controls.<br>No difference in complexity between aMCI and either controls or AD. |
| Li, 2022 [12] | MCI (63), CI (26), controls (32) | Resting state | 1 frontal channel (1S, 2D) | ECO-N17-C25L (Enginmed Bio-Medical Electronics; Suzhou, China) | Reduced TOI in CI compared to controls. |
| Schwarz, 2004 [22] | VaD (1) | Resting state, before/after acupuncture | 2 frontal channels (n/a) | INVOS 5100 (Somanetics; Troy, USA) | Slightly increased regional cerebral oxygen saturation. |
| Liu, 2014 [10] | aMCI (32), controls (21) | Resting state | 1 channel (n/a) | NIRO-200NX (Hamamatsu Photonics CO.; Hamamatsu, Japan) | No difference in TOI between aMCI and controls. |
| Marmarelis, 2017 [9] | aMCI (43), controls (22) | Resting state | 1 frontal channel (n/a) | *Not specified* (Hamamatsu Photonics CO.; Hamamatsu, Japan) | Impaired dynamic $CO_2$ vasomotor reactivity in aMCI compared to controls based on TOI derived from NIRS. |
| Marmarelis, 2021 [18] | aMCI (26), controls (12) | Resting state | 1 frontal channel (n/a) | *Not specified* (Hamamatsu Photonics CO.; Hamamatsu, Japan) | Significant difference in TOI response to change in blood $CO_2$ between MCI and controls. |
| Morimoto, 2022 [14] | Dementia (12) | Resting state, randomised control trial | 1 frontal channel (n/a) | NIRO-200NX (Hamamatsu Photonics CO.; Hamamatsu, Japan) | No difference in TOI between midazolam and dexmedetomidine groups. |
| Nguyen, 2019 [31] * | MCI (42), controls (53) | Resting state | 6 frontal channels (2S, 5D) | Emitting diodes: OE-MV7385-P (Opto ENG; Korea) and photodiodes: Opto101 (Texas Instruments) | Higher Hb connectivity in right frontal cortex in MCI compared to controls. |
| Niu, 2019 [34] | aMCI (25), AD (23), controls (30) | Resting state | 46 whole-head channels (12S, 24D) | CW6 (Techen Co.; MA, USA) | Increased HbO dynamic functional connectivity in aMCI and AD compared to controls. |
| Oyama, 2018 [36] | Severe cognitive impairment (39), MCI (55), controls (108) | Resting state | Frontal channels (n/a) | TRS-21 (Hamamatsu Photonics K.K.; Hamamatsu, Japan) | Significant positive correlation between MMSE score and oxygen saturation measured via time-resolved NIRS. |
| Tarumi, 2014 [11] | aMCI (27), controls (15) | Resting state, during sit/stand procedure | 1 frontal channel (n/a) | NIRO-200X (Hamamatsu Photonics CO.; Hamamatsu, Japan) | Reduced TOI in aMCI compared to controls.<br>Higher TOI correlated with higher behavioural scores in both groups. |

| | | | | | |
|---|---|---|---|---|---|
| Tatsuno, 2021 [14] | Dementia (10), controls (10) | Resting state, before/after midazolam administration | 1 frontal channel (n/a) | NIRO-200X (Hamamatsu Photonics CO.; Hamamatsu, Japan) | No differences in TOI between dementia patients and controls, or upon midazolam administration. |
| Van Beek, 2010 [23] | AD (21), controls (20) | During sit/stand procedure, before/after galantamine treatment | Frontal channels (n/a) | Oxymon (Artinis Medical Systems; Zetten, NL) | Larger decline in AD compared to controls in HbO and Hb upon standing, enhanced with galantamine treatment. |
| Van Beek, 2012 [17] | AD (21), controls (20) | Resting state, during sit/stand procedure | 2 frontal channels (n/a) | Oxymon (Artinis Medical Systems; Zetten, NL) | Larger spontaneous oscillations in HbO in low-frequency range in AD compared to controls. No difference in power spectrum for HbO and Hb for low-frequency and very low-frequency ranges between AD and controls following repeated sit/stand procedures. |
| Viola, 2013 [8] | aMCI (21), controls (10) | Resting state | 4 frontal and temporo-parietal channels (n/a) | T-NIRS EVO II (custom) | Decreased TOI across all ROIs in aMCI compared to controls. |
| Viola, 2014 [13] | Mild AD (10) | Resting state, before/after brain reperfusion rehabilitation therapy | 4 frontal and temporo-parietal channels | T-NIRS EVO II (custom) | Increased TOI in treatment group compared to both the control group and baseline. |
| Yang and Hong, 2021 [33] | MCI (15), controls (9) | Resting state | 48 frontal channels (24S, 32D) | NIRSIT (OBELAB Inc.; Seoul, Korea) | Minimum time window of 30s from resting state measurement of HbO required to detect MCI. |
| Yang, 2022 [85] * | Mild AD (23), MCI (25) | Resting state, before/after physical training | 48 frontal channels (15S, 16D) | NirScan (Danyang Huichuang Medical Equipment Co, Ltd.; Shaanxi, China) | n/a |
| Zeller, 2019 [96] * | MCI (54), controls (61 elderly, 25 young) | Resting state | 52 frontal channels (17S, 16D) | ETG-4000 (Hitachi Medical; Tokyo, Japan) | Reduced power-spectral density of low-frequency oscillations in HbO and Hb in frontal regions in MCI compared to young, but not elderly, controls. |

**Table S2.** Characteristics of the included studies reporting near-infrared spectroscopy data associated with word retrieval in dementia patients and prodromal stages.

| **First author, year** | **Participant characteristics (number)** | **Task design** | **NIRS parameters (number of sources, detectors)** | **NIRS device** | **Main NIRS findings** |
|---|---|---|---|---|---|
| Arai, 2006 [66] | MCI (15), AD (15), controls (32) | Phonemic VFT | 96 frontal, bilateral parietal and occipital channels (n/a) | ETG-7000 (Hitachi Medical; Tokyo, Japan) | Reduced maximum concentration change of HbO in AD in frontal and bilateral parietal areas, and in MCI in right parietal areas, compared to controls. |
| Araki, 2014 [75] | AD (37) | Phonemic VFT, before/after memantine treatment | 22 frontal channels (n/a) | ETG-4000 (Hitatchi Medical; Tokyo, Japan) | Greater change in the value of the HbO integral in treatment group compared to control group in three channels at 24 weeks. |

| | | | | | |
|---|---|---|---|---|---|
| Baik, 2021 [27] * | MCI (22), dementia (20), controls (18) | Semantic VFT | 48 frontal channels (24S, 32D) | NIRSIT (OEBLAB Inc.; Seoul, Korea) | Reduced PFC activity in MCI and dementia group compared to controls, with no difference between dementia and MCI group. |
| Chan, 2020 [30] | Mild AD (16), controls (26) | Semantic VFT | 52 frontal channels (17S, 16D) | OT-R40 (Hitachi Medical; Tokyo, Japan) | Loss of connectivity and insignificant laterality in AD, compared to significant left laterality and higher network efficiency in controls. |
| Fallgatter, 1997 [63] * | AD (10), controls (10) | VFT | 2 frontal channels (2S, 4D) | Critikon 2020 Cerebral Redox Monitor (Johnson and Johnson Medical) | No group difference in average HbR/HBO value.<br>Significant interaction in average HbO value between hemisphere and group. |
| Hermann, 2008 [69] | Dementia (16), controls (16) | VFT | 12 frontal channels (18S, 18D) | ETG-100 (Hitatchi Medical; Tokyo, Japan) | Greater task-related increase in HbO for all channels in controls, and only for some channels in dementia. |
| Ho, 2022 [24] * | Prodromal AD (50), AD dementia (9), asymptomatic AD (28), controls (53) | VFT | 6 frontal channels (2S, 5D) | In house device. | Significant difference in HbO concentration change between groups.<br>Significant difference in HbO concentration change between men and women across patient groups. |
| Hock, 1996 [67] | Probable AD (19), controls (19) | VFT | 4 frontal/parietal channels (n/a) | NIRO 500 (Hamamatsu Photonics CO.; Hamamatsu, Japan) | Decreased HbT and HbO levels in AD compared to controls in the parietal cortex. |
| Hock, 1997 [76] | AD (19), controls (19) | Phonemic VFT | 1 superior parietal channel (n/a) | NIRO 500 (Hamamatsu Photonics CO.; Hamamatsu, Japan) | AD showed decreases in HbO and HbT in parietal cortex, whereas those with mild impairments showed increases in HbO and HbT. |
| Kato, 2017 [68] | AD (42), MCI (65 low-scoring, 33 high-scoring), controls (91) | Shiritori word game | 44 frontal channels (18S, 14D) | ETG-4000 (Hitachi Medical; Tokyo, Japan) | Reduced HbO waveform area and peak amplitude, and increased latency between AD and both controls and high scoring MCI, with no difference compared to low-scoring MCI. |
| Katzorke, 2018 [73] | MCI (55), controls (55) | VFT | 52 fronto-temporal channels (n/a) | ETG-4000 (Hitachi Medical; Tokyo, Japan) | Decreased magnitude of Hb signal in inferior frontotemporal regions in MCI for category VFT compared to controls. |
| Kito, 2014 [74] | AD (28), depressed individuals (30), controls (33) | Phonemic VFT | 44 frontoparietal channels (n/a) | FOIRE-3000 (Shimadzu Corporation; Kyoto, Japan) | No difference in magnitude of HbO signal between AD and controls. |
| Metzger, 2015 [102] | Probable AD (24) | VFT, before/after cholinesterase inhibitor treatment | 44 frontotemporal channels (32S, 28D) | ETG-4000 (Hitachi Medical; Tokyo, Japan) | Increased HbO from baseline in temporal areas with decrease in prefrontal areas. |
| Metzger, 2016 [62] | bvFTD (8), probable AD (8), controls (8) | VFT | 44 frontotemporal channels (32S, 28D) | ETG-4000 (Hitachi Medical; Tokyo, Japan) | Higher activation in frontotemporal regions in controls compared to AD with even more |

| | | | | | |
|---|---|---|---|---|---|
| | | | | | pronounced differences in these regions between bvFTD and controls. |
| Nguyen, 2019 [31] * | MCI (42), controls (53) | VFT | 6 frontal channels (2S, 5D) | Emitting diodes: OE-MV7385-P (Opto ENG; Korea) and photodiodes: Opto101 (Texas Instruments) | Significantly lower connectivity for in HbO and HbT in MCI compared to controls. |
| Richter, 2007 [65] | Dementia (12), controls (12) | VFT, before/after galantamine | 24 frontotemporal channels (18S, 18D) | ETG-100 (Hitachi Medical; Tokyo, Japan) | Reduced haemodynamic response in dementia compared to controls only for female subjects. |
| Yang, 2019 [39] | MCI (15), controls (9) | Semantic VFT | 48 frontal channels (24S, 32D) | NIRSIT (OBELAB Inc.; Seoul, Korea) | Reduced mean HbO change and delayed signal increase in left PFC in MCI compared to controls.<br>No significant difference in mean HbO change for middle or right PFC between MCI and controls. |
| Yang, 2020 [43] * | MCI (15), controls (9) | Semantic VFT | 48 frontal channels (24S, 32D) | NIRSIT (OBELAB Inc.; Seoul, Korea) | Significantly lower mean HbO change in MCI compared to controls. |
| Yap, 2017 [61] | MCI (12), mild AD (18), controls (31) | Semantic VFT | 52 frontotemporal channels (28S, 28D) | OT-R40 (Hitachi Medical; Tokyo, Japan) | No differences between MCI, AD, and controls in mean HbO change.<br>Reduced latency in controls compared to MCI and AD.<br>Steeper slope in controls and MCI compared to AD. |
| Yeung, 2016 [64] | MCI (26), controls (26) | Semantic VFT | 16 frontal channels (6S, 6D) | OEG-$SpO_2$ (Spectratech Inc.; Yokohama, Japan) | No lateralisation of HbO activation in MCI, unlike controls. |
| Yoo, 2020 [40] * | MCI (15), controls (11) | Semantic VFT | 48 frontal channels (24S, 32D) | NIRSIT (OBELAB Inc.; Seoul, Korea) | Reduced magnitude of HbO change in MCI compared to controls. |
| Yoon, 2019 [41] * | MCI (9 aMCI, 6 naMCI), controls (12) | Semantic VFT | 48 frontal channels (24S, 32D) | NIRSIT (OBELAB Inc.; Seoul, Korea) | No differences in accumulated concentration change in HbO across both MCI groups and controls. |

**Table S3.** Characteristics of the included studies reporting near-infrared spectroscopy data associated with memory function in dementia patients and prodromal stages.

| **First author, year** | **Participant characteristics (number)** | **Task design** | **NIRS parameters (number of sources, detectors)** | **NIRS device** | **Main NIRS findings** |
|---|---|---|---|---|---|
| Ates, 2017 [56] | AD (20), controls (20) | Emotional one-back task | 24 frontal channels (n/a) | ETG-4000 (Hitachi Medical; Tokyo, Japan) | Greater change in HbO in left ventral PFC to positive words in AD compared to controls. |
| Baik, 2021 [27] * | MCI (22), dementia (20), controls (18) | N-back task | 48 frontal channels (24S, 32D) | NIRSIT (OEBLAB Inc.; Seoul, Korea) | No difference in PFC activity between groups. |
| Chan, 2021 [53] | MCI (22) | Visual WM task, before/after photobiomodulation | 16 frontal channels (6S, 6D) | OEG-$SpO_2$ (Spectratech Inc.; Yokohama, Japan) | Reduced haemodynamic response in experimental group compared to control group. |

| | | | | | |
|---|---|---|---|---|---|
| Cicalese, 2020 [38] | MCI (8), mild AD (6), controls (8) | Digit encoding and retrieval task | 46 fronto-parietal channels (16S, 15D) | NIRScout (NIRx Medizintechnik GmbH; Berlin, Germany) | Optimal fNIRS features found in the right prefrontal area for classification of dementia stage |
| Ghafoor, 2019 [20] * | MCI (11), controls (11) | Image encoding and retrieval task, before/after acupuncture therapy | 20 frontal channels (8S, 7D) | NIRScout (NIRx Medizintechnik GmbH; Berlin, Germany) | Reduced concentration change of HbO and decreased global efficiency in MCI compared to controls.<br>Increased haemodynamic response with acupuncture therapy in MCI. |
| Ho, 2022 [24] * | Prodromal AD (50), AD dementia (9), asymptomatic AD (28), controls (53) | One-back task | 6 frontal channels (2S, 5D) | In house device. | Significant difference in HbO concentration change between groups.<br>Degree of haemodynamic activation perfectly correlated with AD stage: lowest activation in AD dementia and highest activation in controls. |
| Jang, 2019 [103] | MCI (2) | Delayed matching to sample task, before/after neurofeedback training | 48 frontal channels (24S, 32D) | NIRSIT (OBELAB Inc.; Seoul, Korea) | Increased mean HbO signal after neurofeedback training for one case, which was not present for the other case. |
| Khan, 2022 [52] | MCI (11), controls (11) | Matching to sample task, before/after acupuncture | 20 channels (8S, 8D) | NIRScout (NIRx Medical Technologies; NY, USA) | Significantly greater activation in controls compared to MCI. |
| Kim, 2021 [104] | AD (18), MCI (11), controls (31) | Delayed match to sample task, digit span test | 204 frontal channels (24S, 32D) | NIRSIT (OBELAB Inc.; Seoul, Korea) | HbO changes greater in MCI compared to controls in DMTS. |
| Li, 2018 [49] | aMCI (9), AD (6 mild, 7 severe), controls (8) | Digit verbal span task | 46 fronto-parietal channels (15S, 16D) | NIRScout (NIRx Medizintechnik GmbH; Berlin, Germany) | Reduced HbO change in AD compared to controls in frontal regions.<br>Reduced HbO slope between MCI, mild and severe AD, and controls in frontal regions, with differences only between controls and AD for parietal regions. |
| Li, 2019 [59] | Mild AD (6), controls (8) | Digit verbal span task | 46 fronto-parietal channels (15S, 16D) | NIRScout (NIRx Medizintechnik GmbH; Berlin, Germany) | Higher bilateral symmetry of NIRS activation in controls compared to AD. |
| Li, 2020 [47] | aMCI (16), controls (16) | Digit verbal span task | 30 whole-head channels (16S, 16D) | NIRScout (NIRx Medizintechnik GmbH; Berlin, Germany) | Greater functional connectivity in aMCI compared to controls in several ROIs.<br>Greater global efficiency and clustering coefficients in MCI compared to controls. |
| Nakamura, 2021 [105] | MCI (28), controls (35) | Modified serial number task | 54 whole-head channels (18S, 18D) | LABNIRS (Shimadzu Corporation; Kyoto, Japan) | Novel fNIRS index for quantification of cognitive decline degree showed a significant between-group difference for MCI and controls. |
| Nguyen, 2019 [31] * | MCI (42), controls (53) | One-back task | 6 frontal channels (2S, 5D) | Emitting diodes: OE-MV7385-P (Opto ENG; Korea) and photodiodes: Opto101 (Texas Instruments) | No difference in connectivity between MCI and controls. |

| | | | | | |
|---|---|---|---|---|---|
| Ni, 2021 [48] | Subjective cognitive decline (36) | N-back task, before/after shentai tea polyphenol | 54 whole-head channels (24S, 16D) | Multi-channel NIRS device (Huichang, China) | Increased HbO change following intervention in frontal regions. |
| Niu, 2013 [50] | aMCI (8), controls (16) | Digit n-back tasks | 52 frontal, parietal, and temporal channels (n/a) | ETG-4000 (Hitachi Medical; Tokyo, Japan) | Reduced mean HbO change in aMCI compared to controls in frontal regions. |
| Oboshi, 2016 [58] | AD (11), controls (11) | Visual WM task | 16 frontal channels (6S, 6D) | OEG-16 (Spectratech Inc.; Yokohama, Japan) | Reduced concentration change in HbO in AD compared to controls. |
| Perpetuini, 2017 [57] | AD (11), controls (11) | Free and cued recall task | 17 frontal channels (18S, 8D) | Imagent (ISS Inc.; IL, USA) | Increased signal entropy in Brodmann areas 9 and 46 for early AD compared to controls during delayed free recall. |
| Uemura, 2016 [51] | aMCI (64), controls (66) | Encoding and retrieval task | 22 frontal channels (8S, 7D) | FOIRE-3000 (Shimadzu Corporation; Kyoto, Japan) | Reduced mean HbO in dorsolateral PFC in aMCI compared to controls during retrieval after adjusting for educational history. |
| Ung, 2020 [46] | MCI (12), AD (18), controls (31) | Visuospatial WM task | 52 frontal channels (17S, 16D) | OT-R40 (Hitachi Medical; Tokyo, Japan) | Task load had effect on the slope of haemodynamic response only for MCI. |
| Vermeij, 2017 [45] | MCI (14), controls (21) | N-back task, before/after WM training | 2 frontal channels (n/a) | Oxymon Mk II (Artinis Medical Systems; Einsteinweg, The Netherlands) | Larger mean change in HbR in MCI compared to controls before training.<br>No difference between groups after training. |
| Yang, 2019 [39] | MCI (15), controls (9) | Two-back task | 48 frontal channels (22S, 32D) | NIRSIT (OBELAB Inc.; Seoul Korea) | Increased mean HbO, HbR change, HbO slope, and skewness in the right PFC in MCI compared to controls. |
| Yang, 2020 [43] * | MCI (15), controls (9) | Two-back task | 48 frontal channels (24S, 32D) | NIRSIT (OBELAB Inc.; Seoul, Korea) | Significantly reduced haemodynamic response in MCI compared to controls. |
| Yeung, 2016 [44] * | MCI (26), controls (26) | Digit n-back task | 16 frontal channels (6S, 6D) | OEG-$SpO_2$ (Spectratech Inc.; Yokohama, Japan) | Reduced increases in HbO in high WM load conditions in MCI compared to controls.<br>MCI had smaller increases in frontal activation in response to an increase in WM load compared to controls. |
| Yoo and Hong, 2019 [42] | MCI (15), controls (9) | Two-back task | 48 frontal channels (24S, 32D) | NIRSIT (OBELAB Inc.; Seoul, Korea) | Significantly lower activation (related to HbO signal) in left and right ventrolateral PFC for MCI compared to controls. |
| Yoo, 2020 [40] * | MCI (15), controls (9) | Two-back task | 48 frontal channels (24S, 32D) | NIRSIT (OBELAB Inc.; Seoul, Korea) | Significantly lower activation in left and right ventrolateral PFC for MCI compared to controls |
| Yoon, 2019 [41] * | MCI (9 aMCI, 6 naMCI), controls (12) | Two-back task | (24S, 32D) | NIRSIT (OBELAB Inc.; Seoul, Korea) | No difference in the magnitude of the haemodynamic response between MCI and controls. |
| Yu, 2020 [55] | Cognitive decline (23), controls (23) | Delayed matching to sample task | 120 frontal channels (24S, 32D) | NIRSIT (OBELAB Inc.; Seoul, Korea) | Mean concentration change in HbO was higher for CD compared to controls.<br>Functional connectivity was greater for CD compared to controls. |

**Table S4.** Characteristics of the included studies reporting near-infrared spectroscopy data associated with motor function in dementia patients and prodromal stages.

| First author, year | Participant characteristics (number) | Task design | NIRS parameters (number of sources, detectors) | NIRS device | Main NIRS findings |
|---|---|---|---|---|---|
| Doi, 2013 [78] | MCI (16) | Dual task walking with VFT | 16 frontal channels (6S, 6D) | OEG-16 (Spectratech Inc.; Yokohama, Japan) | Increased mean HbO in dual-task walking compared to normal walking. |
| Nosaka, 2022 [80] | 11 MOCA-J < 26, 14 MOCA-J ≥ 26 | Dual task walking with VFT | 2 frontal channels (3S) | Pocket NIRS HM (Dynasense Co., Ltd.; Wanchai, Hong Kong) | Mean HbO concentration change lower in low cognitive scoring group compared to high scoring group during dual-task walking. |
| Tak, 2011 [84] | Subcortical vascular dementia (6), controls (6) | Hand grip task | 24 fronto-parietal channels (8S, 4D) | Oxymon Mk II (Artinis Medical Systems; Einsteinweg, The Netherlands) | Reduced concentration change of HbO and HbT in SVD patients compared to controls. |
| Talamonti, 2022 [82] | MCI (8), controls (24) | Dual task talking with 2-back task | 256 fronto-motor channels (16S, 16D) | In-house device | MCI showed greater HbO response compared to controls at baseline during single-task walking.<br>No group differences for single or dual-task walking. |
| Teo, 2021 [79] | Subjective memory complaints (23), dementia (9), controls (26) | Dual task walking with number subtraction task | 1 left frontal channel (n/a) | Portalite (Artinis Medical Systems; Zetten, The Netherlands) | Greater increase in HbO in dementia compared to SMC and controls for single-task walking.<br>Increase in HbO in SMC and decrease in HbO in dementia compared to healthy in dual-task walking |
| Wang, 2022 [81] | Cognitive impairment (16), controls (38) | Dual task walking with number subtraction task | 43 whole-head channels (23S, 16D) | Nirsmart (Danyang Huichuang Medical Equipment Co, Ltd.; Shaanxi, China) | No difference in functional connectivity between the two groups during single-task walking.<br>Functional connectivity higher than CI group during dual-task walking. |
| Yang, 2022 [85] * | Mild AD (23), MCI (25) | Finger tapping task | 48 frontal channels (15S, 16D) | NirScan (Danyang Huichuang Medical Equipment Co, Ltd.; Shaanxi, China) | Lower mean HbO concentration change in mild AD compared to MCI. |

**Table S5.** Characteristics of the included studies reporting NIRS data associated with visuo-spatial function in dementia patients and prodromal stages.

| First author, year | Participant characteristics (number) | Task design | NIRS parameters (number of sources, detectors) | NIRS device | Main findings |
|---|---|---|---|---|---|
| Haberstumpf, 2022 [88] | MCI (59), controls (59) | Clock angle discrimination task | 44 parietal channels (16S, 14D) | ETG-4000 (Hitachi Medical; Tokyo, Japan) | Reduced brain activity and laterality in MCI compared to controls. |
| Kito, 2014 [74] * | AD (28), depressed individuals (30), controls (33) | Benton line orientation Task | 22 fronto-parietal channels (n/a) | FOIRE-3000 (Shimadzu Corporation; Kyoto, Japan) | Decreased activation (mean change in HbO) in depressed patients compared to AD. |
| Tomioka et al., 2009 [87] | AD (12), controls (14) | Collision avoidance task | 52 fronto-temporal channels (n/a) | ETG-4000 (Hitachi Medical; Tokyo, Japan) | Increase in HbO concentration was smaller for AD patients compared to controls. |

| | | | | | |
|---|---|---|---|---|---|
| Zeller, 2010 [25] | Probable AD (13), controls (13) | Benton line orientation Task | 24 parietal channels (8S, 8D) | ETG-100 (Hitachi Medical; Tokyo, Japan) | No difference between probable AD and controls for in change in concentration of HbO. |

**Table S6.** Characteristics of the included studies reporting NIRS data associated with other functions in dementia patients and prodromal stages.

| First author, year | Participant characteristics (number) | Task design | NIRS parameters (number of sources, detectors) | NIRS device | Main findings |
|---|---|---|---|---|---|
| Baik, 2021 [27] * | MCI (22), dementia (20), controls (18) | Stroop task | 48 frontal channels (24S, 32D) | NIRSIT (OEBLAB Inc.; Seoul, Korea) | No difference in PFC activity between groups. |
| Fladby, 2004 [92] | Subjective memory complaints, MCI, AD (13 including all patients), controls (8) | Olfactory stimulus | 2 channels over temporal pole and superior temporal gyrus (n/a) | NIRO 300 (Hamamatsu Photonics CO.; Hamamatsu, Japan) | Difference between HbO and Hb concentration change is reduced in patients compared to controls. |
| Ho, 2022 [24] * | Prodromal AD (50), AD dementia (9), asymptomatic AD (28), controls (53) | Oddball task | 6 frontal channels (2S, 5D) | In house device. | Increased HbO concentration change in controls compared to patient groups. |
| Kim, 2022 [91] | AD (16), MCI (26), controls (55) | Olfactory stimulus | 7 frontal channels (n/a) | Fedy Tech (Shenzhen Fedy Technology Co.; Shenzhen, China) | Oxygenation reduced in MCI compared to controls. |
| Liao, 2020 [54] | MCI (34) | Immediate and delayed recall, before\after virtual reality training | 16 frontal channels (n/a) | OEG-16 (Spectratech Inc.; Yokohama, Japan) | Reductions in mean HbO change following training. |
| Nguyen, 2019 [31] * | MCI (42), controls (53) | Oddball task | 6 frontal channels (2S, 5D) | Emitting diodes: OE-MV7385-P (Opto ENG; Korea) and photodiodes: Opto101 (Texas Instruments) | Similar connectivity between the two groups. |
| Tanaka, 2012 [90] | Dementia (1), controls (5) | Music listening | 2 frontal probes (n/a) | OM-220 (Shimadzu Corporation; Kyoto, Japan) | Large activated percent difference at both prefrontal lobes in dementia patients while listening to music.<br>Large activated percent difference at left prefrontal lobe as dementia symptoms worsen. |
| Yang, 2019 [39] * | MCI (15), controls (9) | Stroop task | 48 frontal channels (22S, 32D) | NIRSIT (OBELAB Inc.; Seoul Korea) | Differences across several signal metrics between groups. |
| Yang, 2020 [43] * | MCI (15), controls (9) | Stroop task | 48 frontal channels (24S, 32D) | NIRSIT (OBELAB Inc.; Seoul, Korea) | Average concentration change of HbO in MCI similar to that of controls. |
| Yoo, 2020 [40] * | MCI (15), controls (11) | Stroop task | 48 frontal channels (24S, 32D) | NIRSIT (OBELAB Inc.; Seoul, Korea) | Several channels showed differences in activation between groups. |
| Yoon, 2019 [41] * | MCI (9 aMCI, 6 naMCI), controls (12) | Stroop task | 48 frontal channels (24S, 32D) | NIRSIT (OBELAB Inc.; Seoul, Korea) | Right-sided hyperactivation in naMCI compared to controls. |

| | |
|---|---|
| | Mean accumulated HbO concentration change greatest in naMCI, followed by controls and MCI. |

b. S2: Quality assessment of the included studies

Of the included case-controlled studies, at least half achieved one star across all criteria, with studies performing worse on reporting the selection of their controls and the ascertainment of exposure (Fig S.1). The four included randomised control trials achieved the following scores: 40%, 60%, 80%, and 100%. Of the five observational cohort studies, two scored 50%, two scored 64%, and one scored 86%. One study could not be quality assessed as it was a case report.

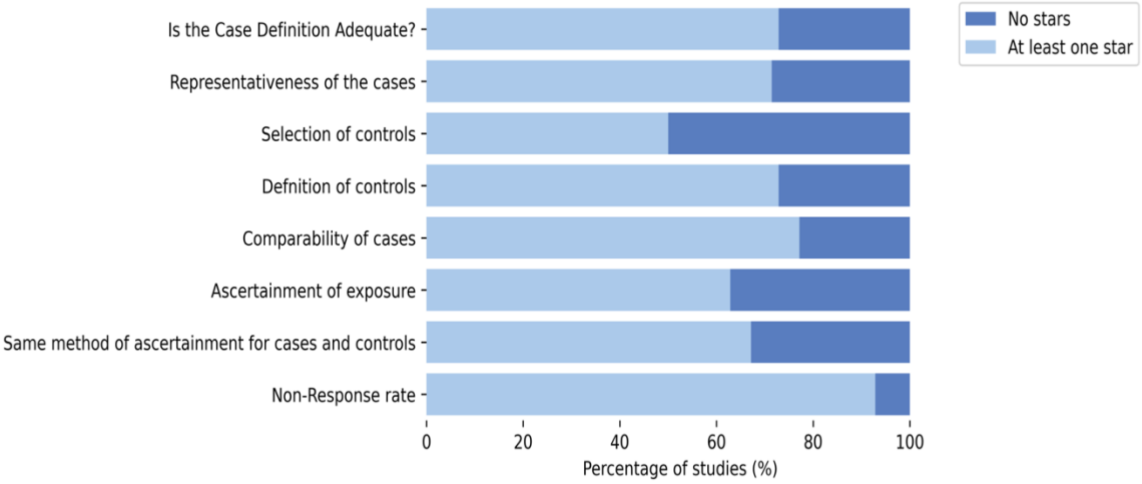


Figure S.1. Quality of the included case-controlled studies as assessed using the Newcastle-Ottawa Scale.
